# Multidimensional Poverty by HIV Status in Eastern and Southern Africa: A Cross-Sectional Analysis of Population-Based HIV Impact Assessment Surveys

**DOI:** 10.64898/2025.12.27.25343083

**Authors:** David Chipanta, Janne Estill, Monica Pinilla-Roncancio, Silas Amo-Agyei, Charles Birungi, Lucas Hertzog, Connie Osborne, Beatrice Matanje, Charles Holmes, Olivia Keiser, Mandeep Dhaliwal

## Abstract

**Introduction:** Multidimensional poverty (deprivations in education, health, and living standards) affects people with and without HIV. We compared poverty levels by HIV status in Eastern and Southern Africa and identified indicators driving deprivations.

**Method:** We analysed the 2020–22 Population HIV Impact Assessment data from Eswatini, Lesotho, Malawi, Mozambique, Tanzania, Uganda, and Zimbabwe using the Alkire-Foster method, calculating the multidimensional poverty index (MPI), headcount ratio, and poverty intensity, using the Stata 14.2 *mpi* command. We classified individuals deprived in 20·0%–33·3% of indicators as vulnerable to poverty, ≥33·3% as poor, and ≥50·0% as severely poor. We estimated the number of people in each poverty category, and decomposed poverty by indicators. Analyses were survey-weighted, disaggregated by sex, residence, and age (15–24 years), and differences by HIV status tested using the Rao–Scott chi-square test (p <0·05).

**Results:** People living with HIV (PLHIV) comprised 7·1% (11·8 million) of the study population (164·9 million). PLHIV were more likely to be vulnerable to poverty, poor, or severely poor than people without HIV. In Eswatini, with the lowest poverty level, PLHIV had higher MPIs (0·248 95% CI [0·239–0·257]) than people without HIV (0·220 [0·215–0·226]). 53·7% [51·8%–55·5%], 99,000, PLHIV were poor compared to 47·5% [46·4%– 48·6%], 266,000, of people without HIV. In Mozambique, with the highest poverty level, the MPIs were similar for people living with and without HIV, but poverty remained higher among PLHIV (70·2% [67·8%– 72·5%], 1·5 million, versus 69·6% [68·6%–70·5%], 10·5 million). The intensity of poverty did not differ across the countries. Education/employment and living standards accounted most for deprivations.

**Interpretation:** Nearly three-quarters of PLHIV in Eastern and Southern Africa experienced multidimensional poverty. Integrating HIV and poverty-reduction efforts, prioritising education, employment, clean energy, water, sanitation, housing, and assets is required. Including HIV indicators in poverty surveys, and research to accelerate joint progress are required.

**Funding:** This study received no external funding

**Research in context:** *Evidence before this study:* We searched PubMed, Google Scholar, reports by UNAIDS, UNDP, World Bank, and other grey literature, using subject headings and keyword terms ("HIV and poverty", "Differences in poverty between people living with HIV and people not living with HIV", "Multidimensional poverty and HIV", "HIV and sanitation", and "Differences between PLHIV and general population in assets") for English-language publications from January 1, 2005, to September 31, 2025. We reviewed 52 articles published in English (Supplement A1). The studies showed that people living with HIV (PLHIV) face socioeconomic disadvantage, including material deprivation, reduced employment, and limited household assets. The studies find as association between poverty among PLHIV with poorer immunologic and virologic response to antiretroviral therapy, lower adherence, and greater comorbidity. They further show inadequate access to safe drinking water, sanitation, and hygiene, increasing diarrhoeal disease and reducing treatment absorption among PLHIV. Other studies find that people living with HIV are deprived in cooking fuels, leading to upper respiratory infections. Most studies used single indicators of poverty or were restricted to individual settings or countries.

*Added value of this study:* This study, to our knowledge, provides the first multi-country assessment of multidimensional poverty by HIV status in Eastern and Southern Africa. We find across countries, that people living with HIV (PLHIV) were more likely to be vulnerable to poverty, poor, or severely poor than people without HIV. Multidimensional poverty among PLHIV was driven by deprivations in education and employment, and deprivations in living standards such as electricity, clean cooking energy, safe drinking water, sanitation, housing, and household assets.

*Implications of all the available evidence:* Despite major gains in HIV treatment and prevention and in national poverty reduction efforts, PLHIV continue to face socioeconomic and infrastructural disadvantage. These deprivations increase PLHIV’s risk to comorbidities and undermine HIV prevention and treatment services, slowing progress toward the Sustainable Development Goals. Integrating HIV responses with poverty reduction and social protection strategies is essential. Incorporating HIV indicators into national poverty monitoring systems and prioritising investment in education, employment, and essential services can accelerate joint progress towards ending AIDS and reducing poverty, in a context of declining external funding.

## Introduction

HIV and poverty are pressing health and development issues in sub–Saharan Africa, the epicentre of the HIV epidemic, where progress in responding to HIV has been remarkable, and where poverty is high (1,2,3). Eastern and Southern Africa, account for 8·7% of the global population, but (21·1 million) half of people living with HIV (PLHIV) and 47·3% of people living in extreme poverty (on $2·15 per day) (1,2,4). Little is known about how gains in the HIV response and poverty reduction reinforce each other in the lives of people living with and without HIV in the region. Understanding whether and how individuals can benefit from progress in the HIV response and poverty reduction simultaneously is urgent. It requires identifying and implementing development accelerators, to amplify progress on multiple fronts including HIV and poverty reduction (5). The United Nations Development Programme (UNDP) defines development accelerators as actions that advance multiple Sustainable Development Goals (SDGs) target outcomes (6). Despite HIV and poverty inequality reinforcing each other (7), few studies have compared poverty levels between people living with and without HIV (8,9,10,11). This information is key in refining policies to integrate HIV and poverty reduction programs as HIV funding and development aid decline.

In Botswana, households with PLHIV had higher rates of household and individual poverty than households without PLHIV (8). In Cambodia and Vietnam, households with PLHIV were more likely to report fewer assets than households without PLHIV (9,10). The Oxford Poverty and Human Development Initiative (OPHI) and the UNDP have conducted multidimensional poverty analysis in over 100 countries (12). In high-income settings like the United States and the United Kingdom, PLHIV are more likely to live at or below the federal poverty line than the general population (13,11). The studies from Botswana, Cambodia and Vietnam were however conducted more than 10 years ago. The prevalence of HIV and poverty have decreased since those studies were completed (1,2). The United States and the United Kingdom have lower HIV burden and poverty rates than Eastern and Southern Africa. The analyses conducted by OPHI and UNDP did not compare poverty levels, between people living with and without HIV. No studies comparing multidimensional poverty between people living with and without HIV across several Eastern and Southern Africa countries have, to our knowledge, been conducted. The findings of this study aim to fill this gap.

In this article, we assessed whether PLHIV experience higher multi-dimensional poverty levels than people without HIV using the latest (2020–2022) population-based HIV impact assessment (PHIV) survey data from Eastern and Southern African. Multidimensional poverty analysis provides a comprehensive picture of poverty in people’s lives spanning health, education, and living standards that income measures alone do not capture (14). We hypothesized that PLHIV are more likely to be vulnerable to poverty, poor or severely poor than people without HIV. PLHIV face stigma, limited access to healthcare, and economic hardship. While effective HIV treatments allow PLHIV to live to old age, PLHIV often experience more health conditions than people without HIV (15,16,17). HIV and poverty can hinder PLHIV’s access to essential services and slow progress towards the SDGs. Identifying these inequalities can inform policies that improve access to healthcare, reinforce social protection programs, and reduce structural barriers disproportionately affecting PLHIV.

## Methods

### Data sources

We utilized data from the publicly available PHIA survey data conducted from 2020 to 2022 in Eswatini, Lesotho, Malawi, Mozambique, Tanzania, Uganda, and Zimbabwe. The PHIA surveys are cross-sectional, household-based, and nationally representative, measuring progress in the 15 U.S. President’s Emergency Plan for AIDS Relief (PEPFAR) supported countries. The surveys focused on HIV among adults aged 15 years and older. Participation in the surveys was voluntary. Data collection, conducted by trained staff, included household and individual interviews, laboratory tests, and the immediate return of HIV test results. The Ministry of Health in each participating country oversaw the surveys, which were funded by the PEPFAR through the U.S. Centers for Disease Control and Prevention, with technical support from the International Center for AIDS Care and Treatment Programs at Columbia University. We combined the Child, Household, Adult, and Adult HIV Biomarker datasets. Detailed information on survey design and sampling can be found through the PHIA Data Manager (https://phia.icap.columbia.edu/).

### Variables and Outcomes

We adapted the structure of the Global multidimensional poverty index (MPI), created in 2010 by OPHI and UNDP, to assess multidimensional poverty. Applied in over 100 countries and included in UNDP Human Development Reports, the MPI guides poverty reduction efforts and tracks SDG progress (14). Following the Global MPI method, we applied the Alkire-Foster Method, using 10 indicators distributed across the domains of education, health and living standards. The sum of the weighted indicators for education, health, and living standards is equal. Within each domain, the weighted indicators add up to one-third.

The Alkire-Foster Method contains three basic statistics: The headcount ratio (H), is the proportion of people who are multidimensionally poor. The intensity of poverty (A) represents the proportion of deprivations among multidimensionally poor individuals. The MPI or the adjusted headcount ratio, a product of headcount ratio and intensity of poverty, reflects the proportion of the population that is multidimensionally poor, accounting for the intensity in poverty. MPI ranges from 0 to 1, whereas the headcount ratio and intensity of poverty ranges from 0 to 100, with higher values reflecting greater poverty.

Other variables we included were sex, age in years grouped into 15-24, 25 -55+, HIV status, education, and rural-urban residence. In Lesotho we combined urban and peri-urban areas to create an urban area variable. We also included HIV viral load suppression because HIV treatment improves the productivity of labour (Supplement Table A2).

### Analysis

The unit of analysis was the individual, classified as living with or without HIV. We assessed deprivation in education/employment, health, and living standards and respective indicators (Figure 1). We proxied the education domain of the Global MPI with education/employment combined. We measured deprivation in education/employment by two indicators at the individual level: not completing secondary education and not having paid employment (cash or kind) in the past year. Similarly, for health, we defined deprivation based on two indicators: the death of any household member in the past five years or the respondent suffering from a chronic condition such as diabetes, hypertension, heart or lung disease, kidney disease, cancer, or a mental health disorder. We measured living standards using six indicators: clean cooking fuel, electricity, safe drinking water, sanitation, housing, and asset ownership (Figure 1).

**Figure 1:**
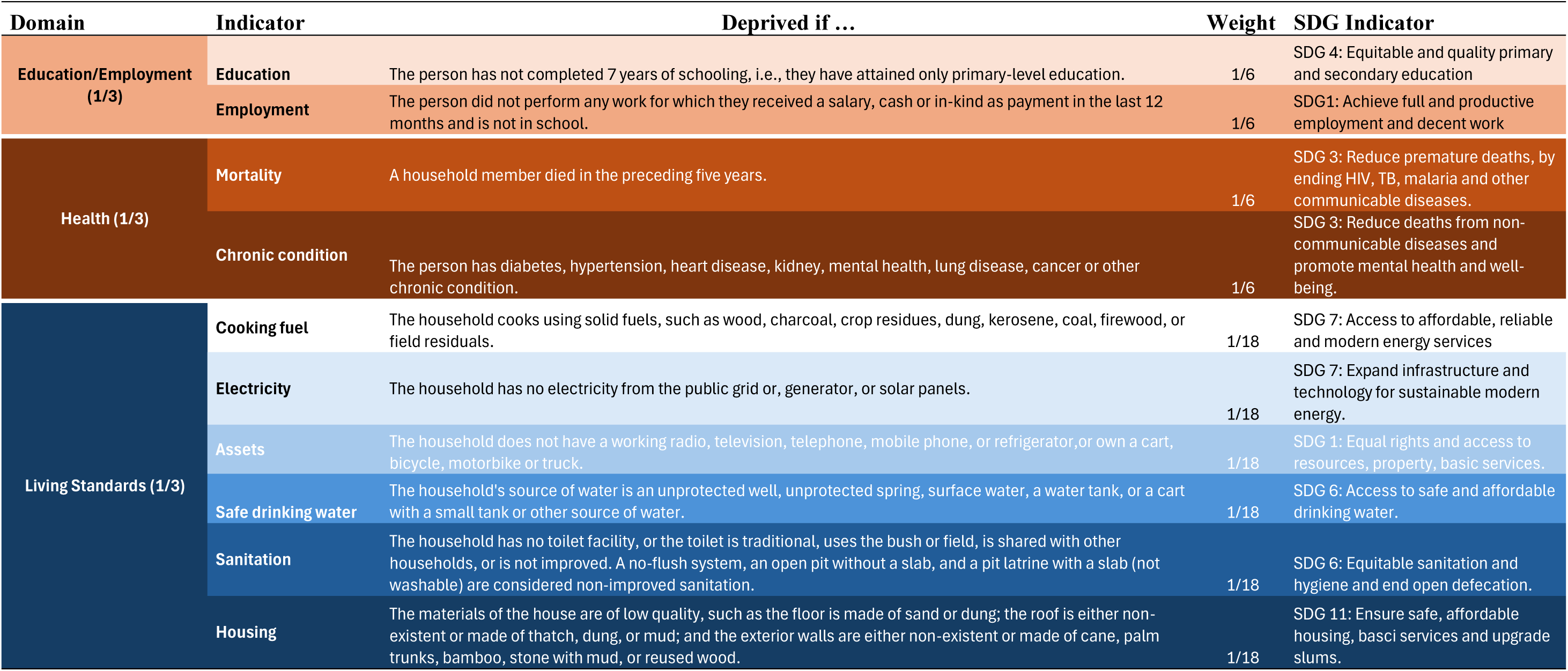
Multiple Poverty Index domain, indicators, deprivation cut-offs and weights. SDG = Sustainable Development Goal

For each country, we described the sample by HIV status, sex, age, education, employment, and rural–urban residence. We set poverty at 33·3% in line with the Global MPI; defining individuals with at least one weighted indicator, as multidimensionally poor. Individuals with deprivations between 20·0%–33·3% were vulnerable to poverty, and those with ≥50·0% were severely poor.

We calculated the headcount ratio, intensity of poverty, their 95% confidence intervals for each cut-off, and the multidimensional poverty index (MPI) using the stata *mpi* command. We also calculated the total number of people who were multidimensionally poor, by multiplying the headcount ratio with the respective population size (14). Supplement A3 shows an example of the calculation of the MPI indices.

To assess differences by HIV status, we computed the relative percentage change for each indicator using the formula:

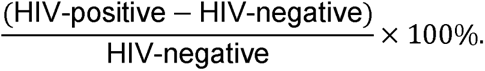

We categorized inequalities as extreme (≥50%), high (20–49%), moderate (5–19%), or low (0–4%).

We further decomposed multidimensional poverty by HIV status across sex, age, and rural–urban residence to examine how each indicator contributed to poverty. These dimensions were selected because they are widely recognized as core components of poverty.

We compared indices proportions by HIV status using the Rao–Scott chi-square test, applying a significance threshold of p < <0·05.

We assessed the MPI’s robustness using the Pearson, Spearman and Kendall rank tests by (a) modifying indicator definitions such as restricting analysis to PLHIV with supressed HIV viral load, redefining mortality as any death in the household in the past year instead of the past five years, limiting chronic conditions to diabetes, hypertension, and heart disease, and excluding charcoal from clean cooking fuel definition; (b) varying poverty cutoffs of 20·0%, 33·3%, and 50·0%; and (c) alternately weighting domains to 50·0% for employment/education, 25·0% each for health, and living standards (18). If the rankings changed significantly, then the MPI would questionable. We used the Stata *mpi* command to calculate indices (19) ‘svy’ command to account for complex sample design (14) and in Stata Version 14.2.

## Results

Across the seven countries, the sample sizes ranged from 11 199 participants in Eswatini to 33 663 in Tanzania, representing 746 000 and 35·3 million people, respectively. A total of 11·8 million people aged 15 years and older were living with HIV, while 153·2 million people in the same age group were not living with HIV (Table 1). The proportion of PLHIV was lowest in Tanzania (4·4% [4·1%–4·7%]) and highest in Eswatini (24·9% [23·7%–26·0%]). Youth living with HIV (aged 15–24) represented a smaller share of the sample of PLHIV, ranging from 6·6% [5·9%–7·5%] in Lesotho to 15·8% [13·5%–18·3%] in Mozambique (Table 1).

**Table 1:**
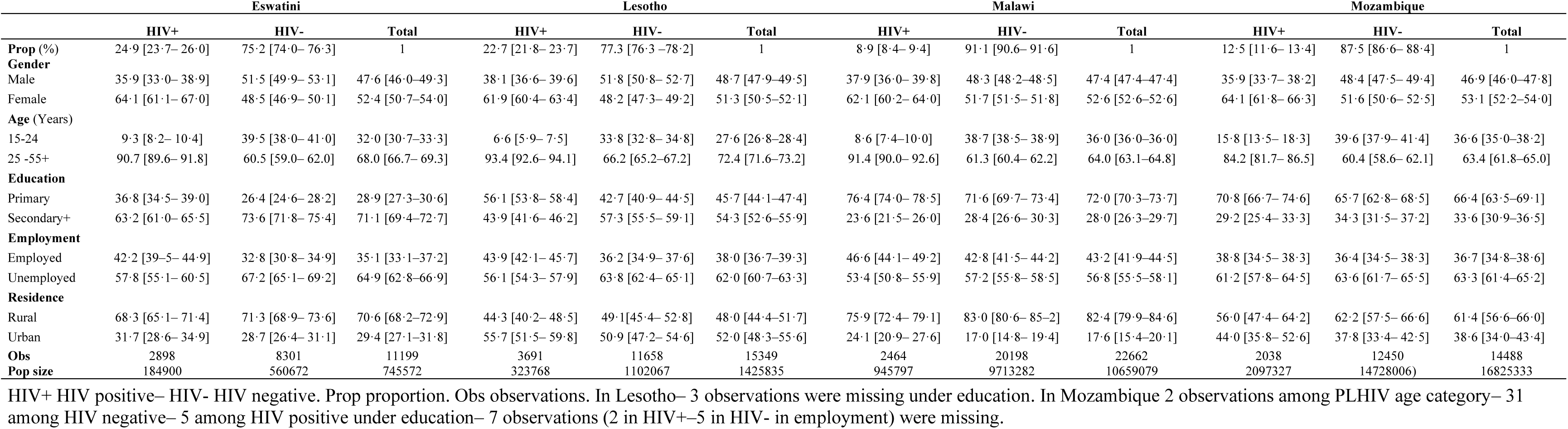
Sample description of characteristics (percent, 95% Confidence Interval, number of observations and population size)

**Table 1:**
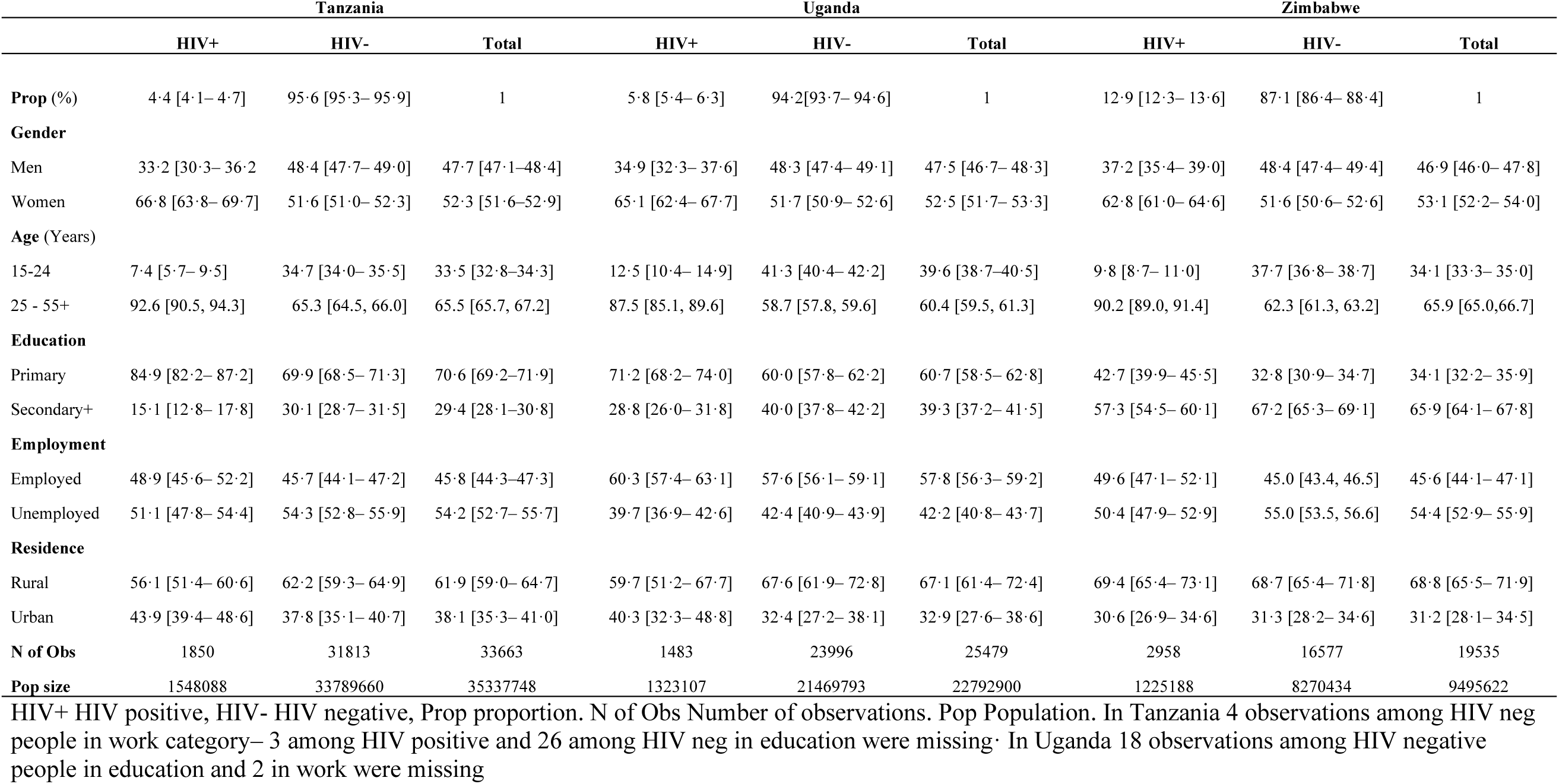
Sample description of characteristics (percent– 95% Confidence Interval– number of observations and population size)

A higher proportion of people living with than without HIV reported living in urban areas, except in Zimbabwe, where we observed no differences, and in Lesotho. A higher proportion of PLHIV tended to have only up to primary level education but were more often employed than people without HIV (Table 1). (Table 1).

In Tanzania and Zimbabwe, PLHIV were more deprived than people without HIV in eight of ten indicators. In the remaining countries PLHIV were more deprived in four to six indicators. We observed the highest levels of deprivation overall in clean cooking fuels, with PLHIV more deprived than people without HIV: 98·9% vs. 98·4% in Malawi (*p* = 0·043), and 93·1% vs. 90·6% in Tanzania (*p* < 0·001), and 98·7% vs. 98·3% in Uganda (*p* = 0·251). Similar for electricity and assets. Employment is the only indicator in which PLHIV were less deprived than people without HIV across all countries surveyed.

The relative inequalities between people living with and without HIV was extreme in chronic conditions in Zimbabwe (137·9%, p < 0·001) and Mozambique (57·0%, p < 0·001), and in education in Eswatini and Lesotho, electricity in Eswatini, and mortality in Lesotho and Malawi. (Figure 2).

**Figure 2:**
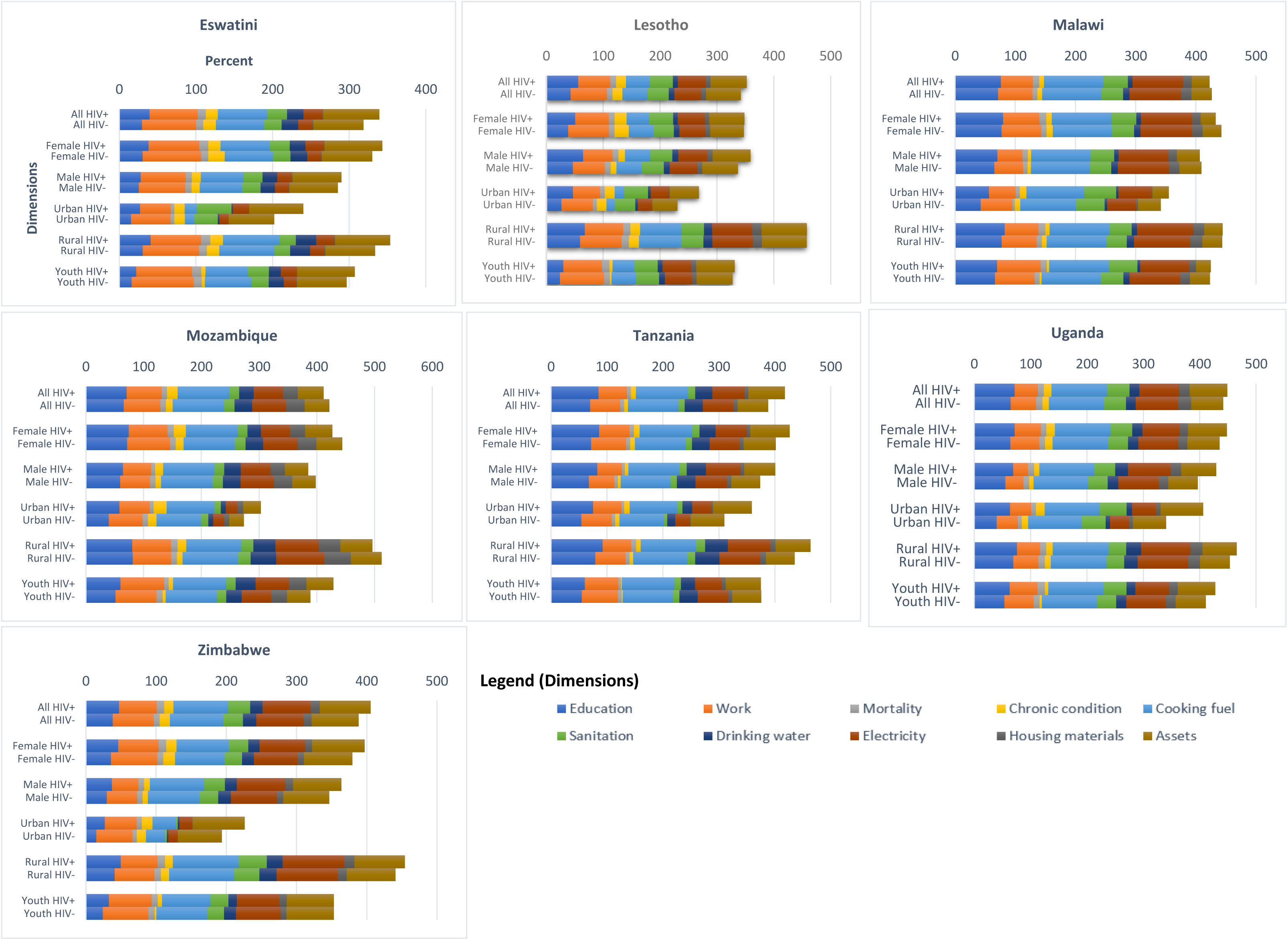
Deprivation in Education/Employment, Health and living standards by HIV status, sex, rural-urban and age.

Across the seven countries an estimated 1.4 [95% CI 1·1–1·7] million PLHIV were vulnerable to poverty; 5·3 [5·2–5·5] million were poor; and 3·5 [3·3–3·6] million were severely poor. Among people without HIV, 17·4 [16·4–18·4] million were vulnerable; 58·8 [58·3–59·4] million were poor; and 37·9 [37·4–38·2] million were severely poor. The MPI comparing people living with and without HIV were: 0·248 [0·239–0·257] versus 0·220 [0·215–0·226]; (all p < 0·01) in Eswatini, the country with the lowest level of poverty. The poverty headcount ratios for people living with and without HIV were 53·7% [51·8%–55·5%] versus 47·5% [46·4%–48·6%]. For Mozambique, which had the highest level of poverty, the corresponding values were 0·350 [0·337– 0·362] and 0·349 [0·344– 0·354] and 70·2 [67·8– 72·5] versus 69·6 [68·6– 70·5]. The intensity of poverty for people living with and without HIV ranged from 46·3 [45·7–46·8] and 46·4 [46·0– 46·8] in Eswatini to 49·8 [49·1– 50·6] and 50·1 [49·8– 50·4] in Mozambique.

Across the dimensions, the disparity between people living with and without HIV was evident in the urban areas of Lesotho, Malawi, Mozambique, Tanzania, and Uganda, and among males in Eswatini and Lesotho, and youth in Malawi. For example, in urban Zimbabwe, the headcount ratio for people living with and without HIV was 28·5 [25·0– 32·0] and 21·3 [20·0– 22·6]; in rural Tanzania it was 84·1 [81·6– 86·5] and 75·0 [74·3– 75·7]. Vulnerability to poverty exceeded 75%, peaking among youth—e.g., in Malawi, 96·8% [94·3%—99·4%] of youth with HIV were vulnerable compared to 91·8% [91·1%—92·4%] without HIV.

The inequality in MPI between people living with and without HIV was small in Lesotho, Malawi, Mozambique, and Uganda, but moderate in Eswatini, Tanzania, and Zimbabwe. Inequalities in headcount ratio were small in Uganda and Mozambique and moderate in Eswatini, Lesotho, Malawi, Tanzania and Zimbabwe. Inequality in poverty intensity was small in the surveyed countries (Table 2).

**Table 2:**
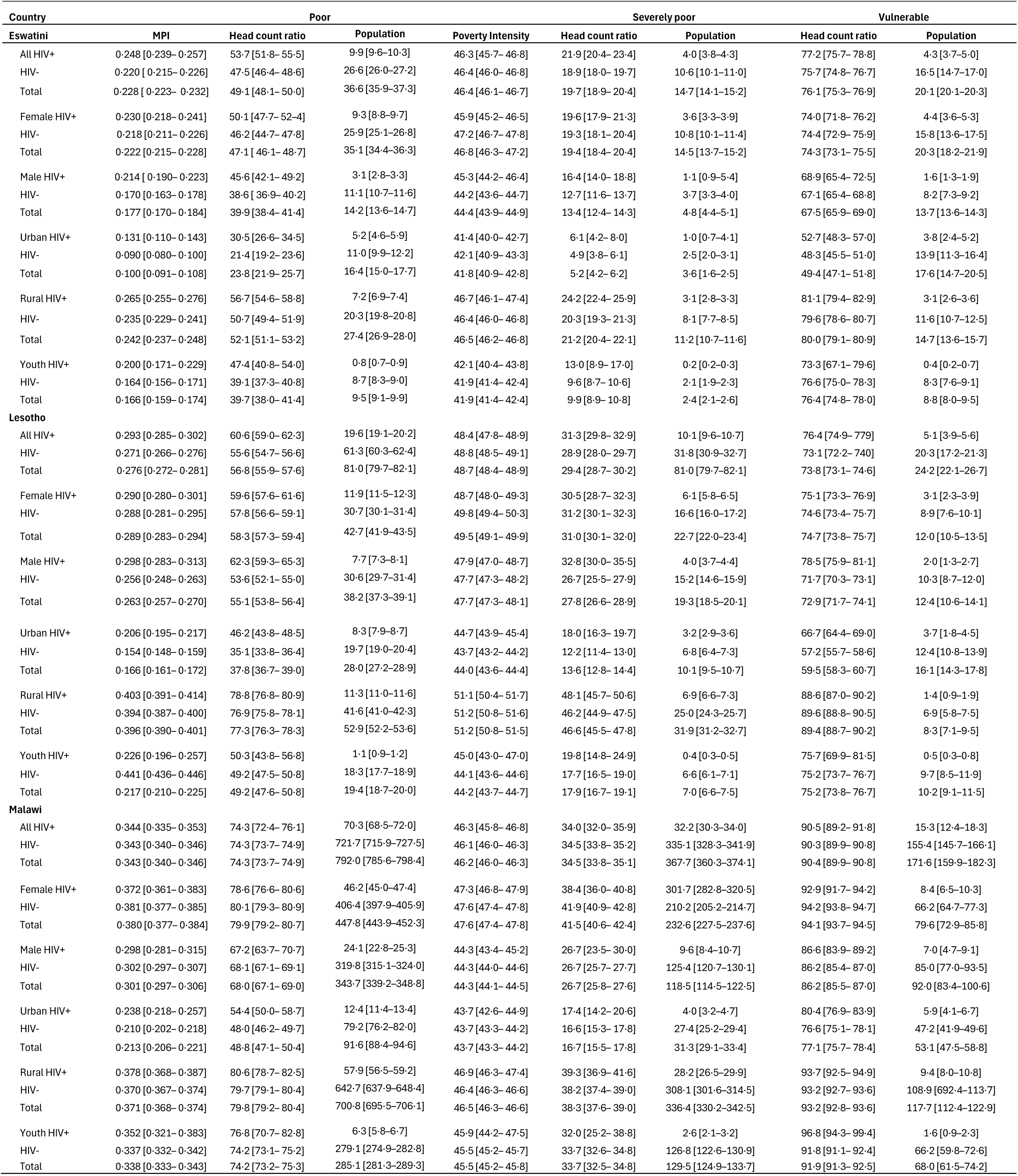
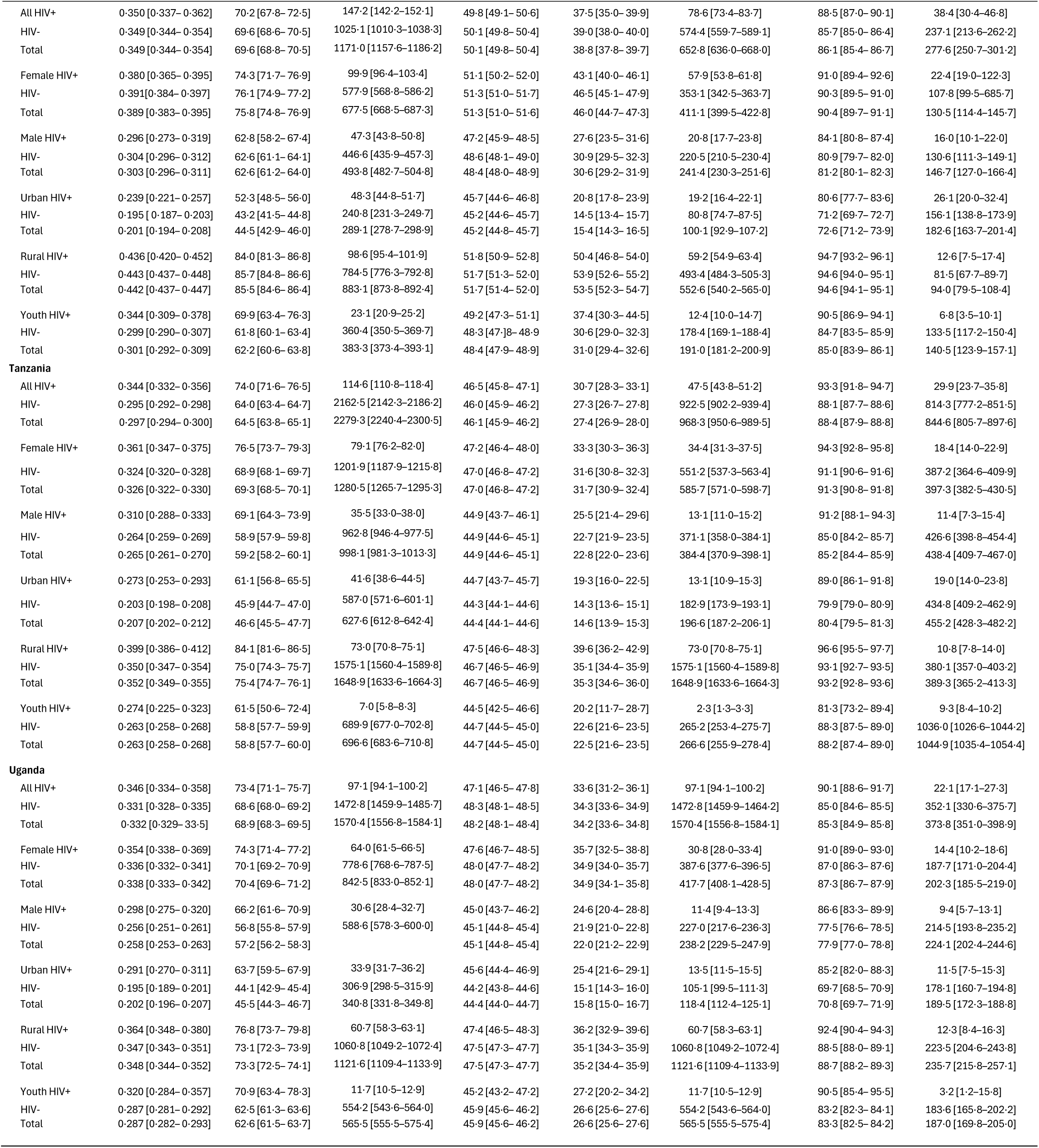
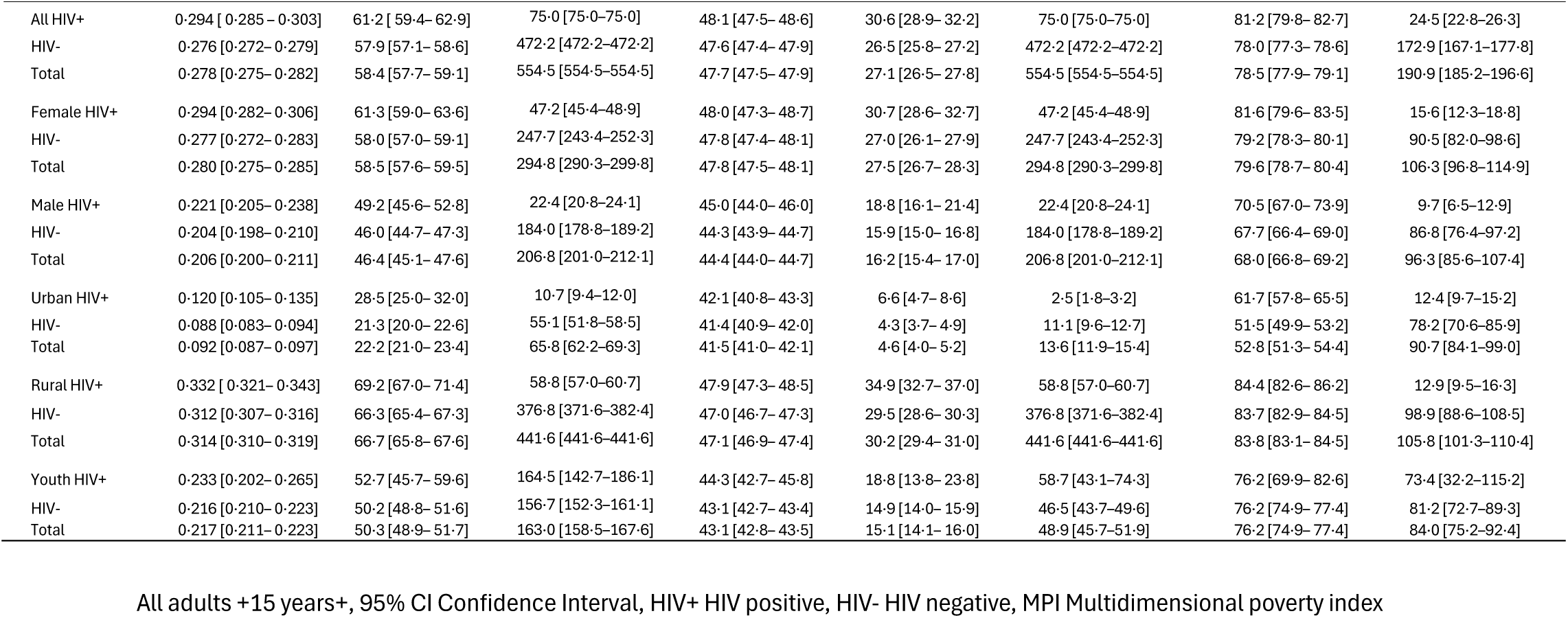
Multidimensional Poverty Indices– Population Proportions Poor, Severely Poor, and Vulnerable [10,000s] by HIV Status and Poverty Dimension [95% CI].

Decomposition analysis showed that education contributed more to poverty among people living with than without HIV in all countries, ranging from 24·2% vs 21·4% in Zimbabwe to 34·1% vs 32·5% (all p < 0·001) in Tanzania. In contrast, employment contributed less to poverty among PLHIV than people without HIV, except in Malawi and Mozambique, where differences were insignificant. Living standards, accounted for 20·0% of poverty in urban Zimbabwe and up to 42·4% among males in Uganda, with similar trends in Lesotho and Tanzania (Figure 2). Health-related deprivations contributed more to poverty among PLHIV in Malawi, Lesotho, and Mozambique, with the highest contribution among youth in Lesotho: 12·1% among youth living with vs 9·2% among youth without HIV, p = 0·037 (Figure 3).

**Figure 3:**
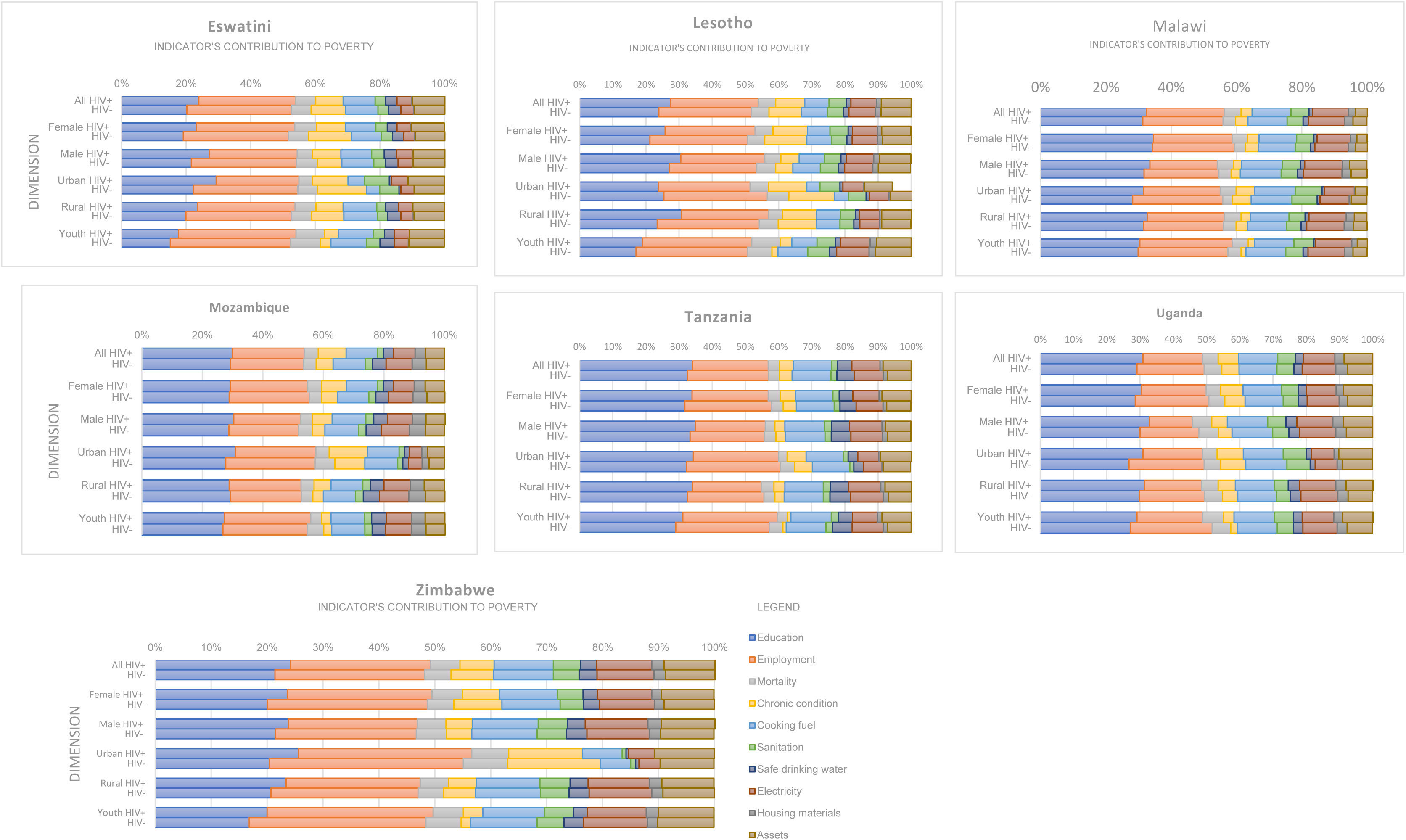
Decomposition of indicators contribution to poverty by HIV status, sex, rural-urban and age.

In terms of robustness of the results, the MPI gaps between people living with and without HIV, and MPI country rankings, remained robust across poverty cutoffs (20·0%–50·0%), alternative weights (50·0%, 25·0%, 25·0%), and indicator adjustments. The correlation coefficients for the Pearson, Spearman and Kendall tau rank tests were large (>0·70) and significant except for the Kendall tau coefficients between indictors weight 1 (50%Education/Employment, 25%Health, 25% Living Standards) and Weight 2 (25%Education/Employment, 50%Health, 25% Living Standards), and poverty cutoffs of 50% and 20% which were 0.62 (*p*= 0·07) (Supplement Table A4).

## Discussion

This study, to our knowledge, is the first to compare multidimensional poverty between people living with and without HIV across seven highly HIV-affected countries in Eastern and Southern Africa. We found, first, that PLHIV were more vulnerable to poverty, more likely to be poor, and often severely poor than people without HIV. In total over 1·3 million PLHIV were vulnerable to poverty, and 5·3 million were poor, of whom 3·4 million were severely poor in the countries included in this study. PLHIV who were youth, men, and living in urban areas were more likely to be poor. Second, PLHIV experienced greater deprivation in up to 8 of 10 indicators with varying levels of relative inequalities. Third, education and living standards contributed more to poverty among people living with than without HIV, while for employment the opposite was true. These findings reveal the critical intersection between HIV-positive status and multidimensional poverty in the environment that shapes people’s health. Policymakers must address the social determinants of health, implement interventions bridging HIV, health and poverty reduction, to accelerate progress in HIV and poverty reduction.

Despite several countries in Eastern and Southern Africa including Eswatini, Lesotho, Malawi, Tanzania, Uganda and Zimbabwe, having achieved near-universal coverage of HIV testing, treatment, and viral suppression (1), PLHIV experienced higher levels of vulnerability, poverty, or severe poverty than people without HIV across the seven countries. The national poverty indices from our study align in general with those from the global MPI, with a few differences (12). The global MPI used nutrition and school attendance indicators from the Demographic Health Survey data sets (12), while we used chronic conditions from the PHIA data sets. These high levels of vulnerability and poverty, and inequality between people living with and without HIV, likely contributed to continued HIV transmission and AIDS-related deaths in the region (20,21,22,23). In 2024, these seven countries had a combined total of 127,900 AIDS related deaths, and 206,400 new HIV infections (1).

The disparity in multidimensional poverty between people living with and without HIV was stark in the urban areas of Lesotho, Malawi, Mozambique, Tanzania, and Uganda; among males in Eswatini and Lesotho; and among youths in Malawi. For example, in Malawi, a significant proportion of youth living with HIV were vulnerable compared to youths without HIV. This result may reflect a high number of children orphaned by HIV in Malawi (1), potentially driving the high level of poverty facing youth in the country. Poverty reduction interventions are required in urban areas and among youths, with a focus on PLHIV. In Uganda, a family-based economic program combining matched savings, mentorship, and financial literacy significantly reduced multidimensional poverty among youths living with HIV (24). Similarly, in sub-Saharan Africa, World Bank aid lowered multidimensional poverty in rich areas of the countries (25). It reduced poverty further when co-located with Chinese aid projects, as Chinese aid helped fill infrastructure gaps (25). Including PLHIV in donor–financed aid may help ensure their inclusion in the systems adopted by Chinese and other infrastructure development funders.

At the indicator level, PLHIV also experienced greater deprivation than people without HIV. In terms of disparities, the relative disparities were pronounced in the burden of chronic conditions among PLHIV. The relative inequalities were extreme in Zimbabwe and high in Mozambique. This result underscores the higher prevalence of noncommunicable diseases among PLHIV even in settings with high ART coverage (26) reflecting higher deprivations in noncommunicable diseases overall. Access to clean cooking fuel was the indicator in which people were most deprived across the countries, followed by access to electricity and assets, with higher deprivation among PLHIV. Countries should emulate the example of Côte d’Ivoire’s near-universal electricity coverage (27). The countries should offer people, including PLHIV from low-income households, flexible financing to afford electricity grid connections alongside improving the generation and supply of electricity (27). Cash transfers paired with clean cooking fuel solutions, such as improved cooking stoves, electric stoves, and biogas reduce indoor air pollution and time spent collecting firewood (28). PLHIV must be engaged in designing and implementing clean cooking energy solutions.

Deprivation in education was common and highest among PLHIV while PLHIV were less deprived in employment across the countries. PLHIV often sell their assets, take up informal jobs and income generating activities to cope with the impact of HIV (29). These activities may be insufficient to improve their living standards. For example, in this study, deprivations in assets were higher among PLHIV than among people without HIV. They ranged from 63·7% for PLHIV and 61·0% (p = 0·004) among people without HIV in Lesotho, to 73·2% and 65·3% (p < 0·0001), respectively, in Eswatini. This result reflects the impact of high HIV prevalence (24·6% and 25·8%, respectively) on poverty. As the largest HIV donor, funder for education, health, social protection and microenterprises and funder of 42% of water and sanitation in low-income countries (30) U.S. funding cuts risk worsening disparities in education/employment, health and living standards between people living with and without HIV. The high multidimensional poverty among PLHIV undermines progress towards achieving SDGs including SDGs 1, 3, 4, 5, 6, 7, and 11. HIV must be integrated and funded within the United Nation 80 reform, ensuring the transition of UNAIDS (31) catalyses a United Nations system-wide drive for HIV-inclusive equity.

The decomposition analysis revealed that education/employment and living standards significantly contributed to poverty among PLHIV than people without HIV across the countries. Living standards deprivation accounted for one-fifth of poverty. Education, jobs, housing, and support need proper funding (32) including for PLHIV. HIV-positive status should be a key criterion for inclusion in national social protection and poverty reduction programs, with eligibility expanded to include more people at risk of poverty (33). Household surveys that measure poverty should include HIV status indicators to monitor and help address the socioeconomic burdens PLHIV face.

This study has several limitations. The cross-sectional nature of the data limits the ability to infer causality between HIV status and multidimensional poverty. Country-level adaptations of the MPI may mask intra-country variations, particularly in conflict and fragile contexts. The data on conflicts and fragility is not included in the PHIA. Limited sample sizes among youth, prevented us from performing sex-disaggregated analyses of poverty by HIV status. Finally, the analysis does not account for the complex interactions among poverty dimensions, domains, and indicators, nor how individual-level deprivation intersects with population-level deprivation. For example, unsafe drinking water affects PLHIV, leading to acute diarrhoea (34) even when access levels are similar to people without HIV. Community sanitation or ambient air quality must improve, often beyond 80% coverage, for individuals to benefit from interventions (35). The value of access to electricity is limited if supply is inconsistent, or unaffordable to households. Despite these limitations, this study is rigorous and generalisable to high HIV burden countries of sub-Saharan Africa.

Finally, we found high levels of poverty with about three quarters of PLHIV experienced multidimensional poverty in Eastern and southern Africa and three quarters to 90% were at risk of falling into poverty including severe poverty. Integrating HIV and poverty-reduction efforts, prioritising education, employment, clean energy, safe drinking water, sanitation, housing, and household assets is required. Research to understand how to achieve joint progress in the HIV response and poverty reduction is required. Household surveys should include HIV indicators to monitor the inclusion of PLHIV in poverty reduction efforts.

## Supporting information

Example of MPI calculations

Variable Description

Ranking Tests

Works consulted

## Data Availability

The data for this study is publicly available and can be requested at PHIA Data Manager (https://phia.icap.columbia.edu/about/).

## Contributions

DC conceptualized the study and led the data curation. DC, MP, and SA defined the methodology. DC performed the data analysis. DC, MP, SA, CB, LC, CO, BM, CH, JE, OK and MD further analysed the data. DC visualized the data. DC drafted the initial manuscript. DC, MP, SA, CB, LC, CO, BM, CH, JE, OK and MD critically reviewed the manuscript. DC further revised the manuscript. All authors saw and verified the data. All authors read and agreed to submit the manuscript.

## Acknowledgements

None.

Appendix Supplementary materials (4)

## References

1. Joint United Nations Programme on HIV/AIDS. AIDSinfo: Global data on HIV epidemiology and response. Geneva: Joint United Nations Programme on HIV/AIDS; 2025. Report No.: www.aidsinfo.unaids.org retrived on September 23, 2025.

2. World Bank. Poverty, Prosperity, and Planet Report 2024: Pathways Out of the Polycrisis. Washington, DC: : World Bank; 2024. Report No.: doi:10.1596/978-1-4648-2123-3..

3. Alkire S, Nogales R, Quinn NN, Suppa N. Global multidimensional poverty and COVID-19: A decade of progress at risk? Social Science & Medicine. 2021; 291: 114457.

4. United Nations Population Fund. World population Dashboard 2025. New York: United Nations Population Fund; 2025. Report No.: https://www.unfpa.org/data/world-population-dashboard Downloaded on 18 August 2025.

5. Sherr L, Haag K, Tomlinson M, Rudgard WE, Skeen S, Meinck F, et al. Understanding accelerators to improve SDG-related outcomes for adolescents-An investigation into the nature and quantum of additive effects of protective factors to guide policy making. PLoS One. 2023 Jan; 18(1: doi: 10.1371/journal.pone.0278020): e0278020.

6. United Nations Development Programme. United Nations Development Programme (Undp). SDG Accelerator and Bottleneck Assessment Tool.. New York: United Nations Development Programme, Sustainable Development Cluster, UNDP Bureau for Policy; 2017.

7. Haacker M, Meyer-Rath G. Interactions Between HIV and Poverty. Policy brief #8 of series “Economic Impact of HIV”. Johannesburg:; 2021.

8. Greener R, Jefferis KR, Siphambe H. The Impact of HIV/AIDS on Poverty and Inequality in Botswana. South African Journal of Economics, Economic Society of South Africa. 2000 December; 68(5)(DOI: 10.1111/j.1813-6982.2000.tb01284.x): 393-404.

9. Marino N, Saint S, C PK, Yasuoka J, Jimba M. Socioeconomic Impact of HIV/AIDS on Households under Free Antiretroviral Therapy in Preah Sihanouk Province, Cambodia. J Antivir Antiretrovir. 2012 December;(DOI: 10.4172/jaa.1000056).

10 Parliamentary Committee for Social Affairs, United Nations Development Programme, Viet Nam. . Impact of HIV/AIDS on household poverty and vulnerabilty in Vietnam. Hai Noi: Parliamentary Committee for Social Affairs, United Nations Development Programme, Viet Nam; 2009.

11. United Kingdom Health Security Agency. Research and analysis. Positive Voices 2022 survey . report. LondoN: Government of the United Kingdom (GOV.UK), United Kingdom Health Security Agency; 2024. Report No.: https://www.gov.uk/government/publications/hiv-positive-voices-survey/positive-voices-2022-survey-report.

12. OPHI (Oxford Poverty and Human Development Initiative) and UNDP (United Nations . Development Programme). Unstacking global poverty: Data for high-impact action. London, New York: OPHI (Oxford Poverty and Human Development Initiative) and UNDP (United Nations Development Programme) , United Nations Development Programme and Oxford Poverty and Human Development Initiative; 2023.

13 Dasgupta S, McManus T, Tie Y, Lin CYC, Yuan X, Sharpe JD, et al. Comparison of Demographic Characteristics and Social Determinants of Health Between Adults With Diagnosed HIV and All Adults in the U.S. AJPM Focus. 2023 September; 2(3: doi.org/10.1016/j.focus.2023.100115): 100115.

14. Alkire S, Kanagaratnam U, Suppa N. The global Multidimensional Poverty Index (MPI) 2024 country results and methodological note’, OPHI MPI Methodological Note 58. London: Oxford Poverty and Human Development Initiative (OPHI), University of Oxford; 2024.

15 Marcus JL, Leyden WA, Alexeeff SE, Anderson AN, Hechter RC, Hu H, et al. Comparison of Overall and Comorbidity-Free Life Expectancy Between Insured Adults With and Without HIV Infection, 2000-2016. JAMA Netw Open. 2020 Jun; 3(6. doi: 10.1001/jamanetworkopen.2020.7954.): e207954.

16 Morales DR, Moreno-Martos D, Matin N, McGettigan P. Health conditions in adults with HIV compared with the general population: A population-based cross-sectional analysis. EClinicalMedicine. 2022 Apr; 47(eCollection 2022 May. doi: 10.1016/j.eclinm.2022.101392): 101392.

17 Payne CF, Houle B, Chinogurei C, Herl MCR, Kabudula CW, Kobayashi LC, et al. Differences in healthy longevity by HIV status and viral load among older South African adults: an observational cohort modelling study. The Lancet HIV. 2022 October; 9(10.): e709 - e716.

18 Alkire S, Kanagaratnam U, Nogales R, Suppa N. Revising the Global Multidimensional Poverty Index: Empirical Insights and Robustness. The review of income and wealth. 2022 March;(10.1111/roiw.12573): S347-S384.

19 Daniele Pacifico FP. Estimating measures of multidimensional poverty with Stata. The Stata Journal. 2017; 17(2): 687–703.

20 Haacker M, Birungi C. Poverty as a barrier to antiretroviral therapy access for people living with HIV/AIDS in Kenya. African Journal of AIDS Research. 2018; 17(2. 10.2989/16085906.2018.1475401): 145–152.

21 Ranganathan M, Heise L, MacPhail C, Stöckl H, Silverwood RJ, Kahn K, et al. ‘It’s because I like things… it’s a status and he buys me airtime’: exploring the role of transactional sex in young women’s consumption patterns in rural South Africa (secondary findings from HPTN 068). Reprod Health. 2018; 15(102. 10.1186/s12978-018-0539-y).

22 Czechowski K, Sylvestre J, Corsini-Munt S. Survival sex: Sexual agency and consent in a state of deprivation? A scoping review. The Canadian Journal of Human Sexuality. 2022 Oct; 31(https://api.semanticscholar.org/CorpusID:251999996): 293 -308.

23. Lépine A, Cust H, Treibich C. What Drives HIV in Africa? Addressing Economic Gender Inequalities to Close the HIV Gender Gap. Oxford Research Encyclopedia of Economics and Finance. 2023 Nov;(https://oxfordre.com/economics/view/10.1093/acrefore/9780190625979.001.0001/acrefore-9780190625979-e-880.).

24 Dvalishvili D, Ssewamala FM, Nabunya P, Bahar OS, Kizito S, Namuwonge F, et al. Impact of Family-Based Economic Empowerment Intervention,Suubi+Adherence (2012–2018) on Multidimensional Poverty for Adolescents Living with HIV (ALWHIV) in Uganda. Int. J. Environ. Res. Public Health. 2022 Sept; 19(Doi.org/10.3390/ijerph192114326): 14326.

25 Zhang L, Li X, Zhuang Y, Li N. World Bank aid and local multidimensional poverty in Sub-Saharan Africa. Economic Modelling. 2022 September; 117(10.1016/j.econmod.2022.106065): 106065.

26 Moyo-Chilufya M, Maluleke K, Kgarosi K, Muyoyeta M, Hongoro C, Musekiwa A. The burden of non-communicable diseases among people living with HIV in Sub-Saharan Africa: a systematic review and meta-analysis. EClinicalMedicine. 2023 Oct; 65(doi: 10.1016/j.eclinm.2023.102255): 102255.

27. International Finance Cooperation WBG. Côte d’Ivoire Nears Universal Access to Electricity.. Report No.: https://www.ifc.org/en/stories/2025/cote-divoire-nears-universal-access-to-electricity downloaded on September 29, 2025.

28 Chakrabarti A, Handa S, Teams obotMaZCTE. The impacts of cash transfers on household energy choices. American journal of agricultural economics. 2023 January;(10.1111/ajae.12366): 1426-1457.

29 Fauk NK, Mwakinyali SE, Putra S, Mwanri L. Understanding the strategies employed to cope with increased numbers of AIDS-orphaned children in families in rural settings: a case of Mbeya Rural District, Tanzania. Infect Dis Poverty. 2017; 21(10.1186/s40249-016-0233-7): 6.

30 Cavalcanti DM, Sales LdOFd, Silva AFd, Basterra EL, Pena D, Monti C, et al. Evaluating the impact of two decades of USAID interventions and projecting the effects of defunding on mortality up to 2030: a retrospective impact evaluation and forecasting analysis. The Lancet. 2025 Jul; 406(10500. 10.1016/S0140-6736(25)01186-9): 283 - 294.

31 Nations U. UN80 Initiative. Shifting paradigms. United to deliver. Report of the Secretary General. New York: United Nations; 2025.

32 Marmot M. Society and the slow burn of inequality. The Lancet. 2020 May; 395(10234): 1413–1414.

33 Chipanta D, Amo-Agyei S, Hertzog L, Hosseinpoor AR, Smith M, Mahoney C, et al. Missing the vulnerable—Inequalities in social protection in 13 sub-Saharan African countries: Analysis of population-based surveys. Plos Global Health. 2024 July;(10.1371/journal.pgph.0002973).

34 Yates T, Lantagne D, Mintz E, Quick R. The Impact of Water, Sanitation, and Hygiene Interventions on the Health and Well-Being of People Living With HIV. JAIDS Journal of Acquired Immune Deficiency Syndromes. 2015 April; 68(DOI:10.1097/QAI.0000000000000487): S318-S330.

35 Berkouwer SB, Dean JT. Cooking, Health, and Daily Exposure to Pollution Spikes. AMERICAN ECONOMIC JOURNAL: ECONOMIC POLICY. 2025 March;(SSN 1945-774X (Online) DOI 10.3386/w31614).

