## Supplementary material for "Multidimensional Poverty by HIV Status in Eastern and Southern Africa: A Cross-Sectional Analysis of Population-Based HIV Impact Assessment Surveys": Example of MPI calculations

### Supplementary A 3. Detailed Methods Explanation with Example Calculations

#### 1. Overview of the Alkire–Foster MPI Framework

The Alkire–Foster (AF) method is used to measure multidimensional poverty. It evaluates individuals across multiple indicators covering education, health, and living standards. Each person’s deprivation status is compared to predefined thresholds: Vulnerable (20·0–33·3%), MPI poor (≥33·3%), Severely poor (≥50·0%). Selected indicators are assigned equal weights (10 indicators × 0·1 each). Also, a poverty cutoff, is selected denoted by $k$. In the global Multidimensional Poverty Index (MPI), $k=\frac{1}{3}$.

#### 2. Steps in the Alkire–Foster Method

Suppose we measure people’s living conditions using d different indicators (for example, education, health, or housing).
For each indicator:

- $x_{ij}$is person *i*’s achievement in indicator *j*
- $z_{j}$is the cutoff that determines whether someone is deprived in indicator *j*
- $w_{j}$is the weight given to indicator *j*, where all weights add up to 1

**Step 1: Individual deprivation score**

A person is considered deprived in an indicator if their achievement falls below the cutoff for that indicator.

The deprivation score for person *i* is calculated by:

- Checking each indicator to see whether the person is deprived
- Adding up the weights of all indicators in which they are deprived

Formally, the deprivation score is: $c_{i}=\sum_{j=1}^{d} w_{j}\text{ }I(x_{ij}<z_{j})$

where the indicator function $I(\cdot)$equals 1 if the person is deprived and 0 otherwise.

So, $c_{i}$represents the share of weighted indicators in which person *i* is deprived.

**Step 2: Identifying who is poor**

- A person is considered **multidimensionally poor** if their deprivation score is at least $k$: $c_{i}\geq k$
  To focus only on the poor, we define the **censored deprivation score**: $c_{i}(k)=c_{i}\text{ }I(c_{i}\geq k)$

This means: Poor individuals keep their deprivation score, non-poor individuals are assigned a score of zero

**Step 3: Multidimensional Poverty Index (MPI)**

The overall level of multidimensional poverty in a population is the average censored deprivation score:

$$M=E(c_{i}(k))$$

This is called the MPI.

**Step 4: Breaking MPI into incidence and intensity**

Using probability rules, the MPI can be rewritten as: $MPI=A\times H$
where: $H$is the headcount ratio: the proportion of people who are multidimensionally poor $H=P(c_{i}\geq k)$
$A$is the intensity of poverty: the average deprivation score among the poor only $A=E(c_{i}\mid c_{i}\geq k)$

Interpretation: $H$tells us **how many people are poor.** $A$tells us **how poor they are on average.** $MPI$combines both pieces of information. For this reason, the MPI is often described as an **adjusted headcount ratio**: it adjusts the share of poor people by how many deprivations they suffer.

##### 3. Example: Deprivation Matrix

Person B: deprived in 3/10 indicators → c_B = 0·3 (vulnerable)
Person C: deprived in 4/10 indicators → c_C = 0·4 (MPI poor)

#### 4. Example Calculation

Five individuals with deprivation scores:

A: 0·10 | B: 0·30 | C: 0·40 | D: 0·60 | E: 0·20

MPI poor = individuals with c_i ≥ 0·333 → C and D are poor.

Headcount ratio (H) = 2/5 = 0·4

Intensity (A) = (0·40 + 0.60)/2 = 0·50

Adjusted headcount ratio (M₀ = H × A) = 0·4 × 0·5 = 0·20

Interpretation: 40·0% of MPI are poor and, on average, deprived in 50·0% of indicators (MPI = 0·20).

#### 5. Estimating number of people in each poverty category

To estimate total counts: Number of MPI poor = H × Population.
Example: H = 0·40, Population = 1,000,000 → 0·40 × 1,000,000 = 400,000 MPI poor individuals.

#### 6. Decomposition of Poverty

The MPI can be decomposed by indicator and by group (sex, residence, age). Contribution of indicator j = (w_j × H_j) / MPI, where H_j = proportion of MPI poor deprived in indicator j.
