## Supplementary material for "Multidimensional Poverty by HIV Status in Eastern and Southern Africa: A Cross-Sectional Analysis of Population-Based HIV Impact Assessment Surveys": Variable Description

Supplementary information Table A2. Variable descriptions.

| Variable name | Description | Measure |
| --- | --- | --- |
| HIV status | People 15 years or older testing HIV negative or positive using laboratory confirmed HIV test results from the adult biomarker questionnaire. | Binary |
| Sex | Self-reported sex (i.e., male, female), from the adult interview questionnaire. | Binary |
| Age | Self-reported linear years categorised in years (15–24, 25–54+ and 55 and above), from the adult interview questionnaire. Respondents aged 15-24 years old were defined as youth | Categorical |
| Residency | Self-reported residence (i.e., rural, urban), from the household questionnaire. In Lesotho we combined urban and peri-urban variables to create an urban area variable for consistency in analysis | Binary |
| Employment status | Combined reported working in the past 12 months "Have you done any work in the last 12 months for which you received a salary, cash, or in kind as payment?" from the adult interview questionnaire. | Binary |
| HIV viral load | Combined results of viral load as a continuous variable and categorised <5000 copies/ml as Low HIV viral Load, ≥5000 but ≤15,000 copies/ml, medium, and >15,000 copies/ml as high from the adult biomarker questionnaire. | Categorical |
| Educational status | Level of school that respondent had ever attended (i.e., not educated, primary school education combined into upto primary level education. Secondary, higher education combined to more than secondary), from the adult interview questionnaire. | Categorical |
