## Supplementary material for "Multidimensional Poverty by HIV Status in Eastern and Southern Africa: A Cross-Sectional Analysis of Population-Based HIV Impact Assessment Surveys": Ranking Tests

|  |  | Indicator modifications | | |
| --- | --- | --- | --- | --- |
| Indicator | Test | No change | Mortality in one year | Limited chronic conditions |
| Mortality in one year | Pearson | 0·964 | 1·000 |  |
|  | Spearman | 0·964 |  |  |
|  | Kendall tau b | 0·905 |  |  |
| Limited chronic conditions | Pearson | 0·857 | 0·929 |  |
|  | Spearman | 0·857 | 0·929 |  |
|  | Kendall tau b | 0·714 | 0·810 |  |
| Charcoal off clean cooking energy | Pearson | 0·898 | 0·943 | 0·943 |
|  | Spearman | 0·901 | 0·937 | 0·937 |
|  | Kendall tau b | 0·762 | 0·857 | 0·857 |

|  |  | | Cutt-Off | | | | | |
| --- | --- | --- | --- | --- | --- | --- | --- | --- |
| Cutt-off | Test | | 33·30% | | 20·0% | | 50·% | |
| 20% | Pearson | | 1·000 | |  | |  | |
|  | Spearman | | 1·000 | |  | |  | |
|  | Kendall tau b | | 1·000 | |  | |  | |
| 50% | Pearson | | 0·714 | | 0·714 | |  | |
|  | Spearman | | 0·714 | | 0·714 | |  | |
|  | Kendall tau b | | 0·619 | | 0·619 | |  | |
| 33.3% Viral Load Suppression | Pearson | | 1·000 | | 1·000 | | 0·714 | |
|  | Spearman | | 1·000 | | 1·000 | | 0·714 | |
|  | Kendall tau b | | 1·000 | | 1·000 | | 0·619 | |
|  | |  | | Domain Weights | | | | |
| Domain Weights | | Tests | | Equal | | Weight 1: | | Weight 2 |
| Weight 1: | | Pearson | | 0·964 | |  | |  |
|  |  | Spearman | | 0·964 | |  | |  |
|  |  | Kendall tau b | | 0·905 | |  | |  |
| Weight 2: | | Pearson | | 0.857 | | 0·821 | |  |
|  |  | Spearman | | 0·857 | | 0·821 | |  |
|  |  | Kendal tau b | | 0·714 | | 0·619 | |  |
| Weight 3: | | Pearson | | 0·964 | | 0·929 | | 0·893 |
|  |  | Spearman | | 0·964 | | 0·929 | | 0·893 |
|  |  | Kendall tau b | | 0·905 | | 0·810 | | 0·810 |

Legend

Equal: Education/Employment, Health, Living Standards.

Weight 1: 50·0%Education/Employment, 25·0%Health, 25·0% Living Standards

Weight 2: 25·0%Education/Employment, 50·0%Health, 25·0% Living Standards.

Weight 3: 25·0%Education/Employment, 25·0%Health, 50·0% Living Standards.

Number of countries = 7
