## Supplementary material for "Multidimensional Poverty by HIV Status in Eastern and Southern Africa: A Cross-Sectional Analysis of Population-Based HIV Impact Assessment Surveys": Works consulted

Supplementary Table 1: Works consulted

1. Alkire S, Kanagaratnam U, Nogales R, Suppa N. Revising the Global Multidimensional Poverty Index: empirical insights and robustness. Rev Income Wealth. 2022 Mar;68(Suppl 1):S347–S384. doi:10.1111/roiw.12573.
2. Alkire S, Kanagaratnam U, Suppa N. The Global Multidimensional Poverty Index (MPI) 2024: country results and methodological note. OPHI MPI Methodological Note 58. Oxford: OPHI, University of Oxford; 2024.
3. Alkire S, Nogales R, Quinn NN, Suppa N. Global multidimensional poverty and COVID-19: a decade of progress at risk? Soc Sci Med. 2021;291:114457.
4. Ayer A, Mukondwa RW, Avilés-Guamán C, Takarinda K, West N, Makoni T, et al. Poverty and protection: the relationship between multidimensional poverty, social protection interventions, and HIV viral load. AIDS. 2025 Nov;39(13):1926–1935. doi:10.1097/QAD.0000000000004276.
5. Berkouwer SB, Dean JT. Cooking, health, and daily exposure to pollution spikes. Am Econ J Econ Policy. 2025 Mar. doi:10.3386/w31614.
6. Cavalcanti DM, de Oliveira Ferreira de Sales L, da Silva AF, Basterra EL, Pena D, Monti C, Barreix G, Silva NJ, Vaz P, Saute F, Fanjul G, Bassat Q, Naniche D, Macinko J, Rasella D. Evaluating the impact of two decades of USAID interventions and projecting the effects of defunding on mortality up to 2030: a retrospective impact evaluation and forecasting analysis. Lancet. 2025 Jul 19;406(10500):283-294. doi: 10.1016/S0140-6736(25)01186-9. Epub 2025 Jun 30. PMID: 40609560; PMCID: PMC12274115.
7. Chakrabarti A, Handa S. The impacts of cash transfers on household energy choices. Am J Agric Econ. 2023 Jan;105:1426–1457. doi:10.1111/ajae.12366.
8. Chipanta D, Amo-Agyei S, Hertzog L, Hosseinpoor AR, Smith M, Mahoney C, et al. Missing the vulnerable: inequalities in social protection in 13 sub-Saharan African countries. PLoS Glob Public Health. 2024;4:e0002973. doi:10.1371/journal.pgph.0002973.
9. Chipanta D, Estill J, Stöckl H, Hertzog L, Toska E, Chanda P, Mwanza J, Kaila K, Matome C, Tembo G, Keiser O, Cluver L. Associations of Sustainable Development Goals Accelerators With Adolescents' Well-Being According to Head-of-Household's Disability Status-A Cross-Sectional Study From Zambia. Int J Public Health. 2022 Feb 25;67:1604341. doi: 10.3389/ijph.2022.1604341. PMID: 35283719; PMCID: PMC8916123.
10. Chipanta D, Kapambwe S, Nyondo-Mipando AL, Pascoe M, Amo-Agyei S, Bohlius J, Estill J, Keiser O. Socioeconomic inequalities in cervical precancer screening among women in Ethiopia, Malawi, Rwanda, Tanzania, Zambia and Zimbabwe. BMJ Open. 2023 Jun 20;13(6):e067948. doi:10.1136/bmjopen-2022-067948.
11. Czechowski K, Sylvestre J, Corsini-Munt S. Survival sex: sexual agency and consent in a state of deprivation? Can J Hum Sex. 2022 Oct;31:293–308.
12. Dasgupta S, McManus T, Tie Y, Lin CYC, Yuan X, Sharpe JD, et al. Comparison of demographic characteristics and social determinants of health between adults with diagnosed HIV and all adults in the United States. AJPM Focus. 2023 Sep;2(3):100115. doi:10.1016/j.focus.2023.100115.
13. Dinçer M, Köse N, Ünal E. The role of socioeconomic and behavioral factors in HIV-related deaths. Humanit Soc Sci Commun. 2024;11:1588. doi:10.1057/s41599-024-01924-3.
14. Dvalishvili D, Ssewamala FM, Nabunya P, Bahar OS, Kizito S, Namuwonge F, et al. Impact of the Suubi+Adherence economic empowerment intervention on multidimensional poverty among adolescents living with HIV in Uganda. Int J Environ Res Public Health. 2022 Sep;19:14326. doi:10.3390/ijerph192114326.
15. Fauk NK, Mwakinyali SE, Putra S, Mwanri L. Understanding the strategies employed to cope with increased numbers of AIDS-orphaned children in families in rural Tanzania. Infect Dis Poverty. 2017;6:21. doi:10.1186/s40249-016-0233-7.
16. Greener R, Jefferis KR, Siphambe H. The impact of HIV/AIDS on poverty and inequality in Botswana. S Afr J Econ. 2000 Dec;68(5):393–404. doi:10.1111/j.1813-6982.2000.tb01284.x.
17. Haacker M, Birungi C. Poverty as a barrier to antiretroviral therapy access for people living with HIV/AIDS in Kenya. Afr J AIDS Res. 2018;17(2):145–152. doi:10.2989/16085906.2018.1475401.
18. Haacker M, Meyer-Rath G. Interactions between HIV and poverty. Johannesburg: Policy Brief; 2021.
19. Hasell J, Arriagada P, Rohenkohl B. Beyond income: understanding poverty through the Multidimensional Poverty Index [Internet]. Our World in Data; 2024 [cited 2025 Nov 11]. Available from: https://ourworldindata.org/multidimensional-poverty-index
20. Ibrahim F, Anderson J, Bukutu C, Elford J. Social and economic hardship among people living with HIV in London. HIV Med. 2008 Aug;9(8):616–624. doi:10.1111/j.1468-1293.2008.00605.x.
21. International Finance Corporation (World Bank Group). Côte d’Ivoire nears universal access to electricity. Washington (DC): IFC; 2025.
22. Ismail SM, Kari F, Kamarulzaman A. The socioeconomic implications among people living with HIV/AIDS in Sudan: challenges and coping strategies. J Int Assoc Provid AIDS Care. 2017;16(5):446–454.
23. Joint United Nations Programme on HIV/AIDS (UNAIDS). AIDSinfo: Global data on HIV epidemiology and response. Geneva: UNAIDS; 2025.
24. Kalichman SC, Hernandez D, Kegler C, Cherry C, Kalichman MO, Grebler T. Dimensions of poverty and health outcomes among people living with HIV infection: limited resources and competing needs. J Community Health. 2015 Aug;40(4):702–708. doi:10.1007/s10900-014-9988-6.
25. Lépine A, Cust H, Treibich C. What drives HIV in Africa? Addressing economic gender inequalities to close the HIV gender gap. Oxford Res Encycl Econ Finance. 2023 Nov.
26. Little TM, Handa S, et al. Accelerators to reduce violence, HIV risk, and early pregnancy among adolescents and young people in Namibia. PLoS Glob Public Health. 2025 May;5(5):e0004633. doi:10.1371/journal.pgph.0004633.
27. Lungu EA, et al. Multidimensional poverty and HIV in Malawi: a nationwide analysis. BMC Public Health. 2019;19(1):1616.
28. Marcus JL, Leyden WA, Alexeeff SE, Anderson AN, Hechter RC, Hu H, et al. Comparison of overall and comorbidity-free life expectancy between insured adults with and without HIV infection, 2000–2016. JAMA Netw Open. 2020 Jun;3(6):e207954. doi:10.1001/jamanetworkopen.2020.7954.
29. Marino N, Saint S, PK C, Yasuoka J, Jimba M. Socioeconomic impact of HIV/AIDS on households under free antiretroviral therapy in Preah Sihanouk Province, Cambodia. J Antivir Antiretrovir. 2012 Dec. doi:10.4172/jaa.1000056.
30. Marmot M. Society and the slow burn of inequality. Lancet. 2020 May;395(10234):1413–1414.
31. McClean AR, Trigg J, Ye M, McLinden T, Kooij KW, Bacani N, et al; CANOC Collaboration. Neighbourhood-level material deprivation and response to combination antiretroviral therapy in the Canadian Observational Cohort (CANOC). CMAJ Open. 2022 Mar 15;10(1):E183–E189. doi:10.9778/cmajo.20200249.
32. Morales DR, Moreno-Martos D, Matin N, McGettigan P. Health conditions in adults with HIV compared with the general population. EClinicalMedicine. 2022 Apr;47:101392. doi:10.1016/j.eclinm.2022.101392.
33. Moyo-Chilufya M, Maluleke K, Kgarosi K, Muyoyeta M, Hongoro C, Musekiwa A. The burden of non-communicable diseases among people living with HIV in sub-Saharan Africa. EClinicalMedicine. 2023 Oct;65:102255. doi:10.1016/j.eclinm.2023.102255.
34. North CM, Valeri L, Hunt PW, Mocello AR, Martin JN, Boum Y II, et al. Cooking fuel and respiratory symptoms among people living with HIV in rural Uganda. ERJ Open Res. 2017;3(2):00094–2016. doi:10.1183/23120541.00094-2016.
35. Oxford Poverty and Human Development Initiative (OPHI), United Nations Development Programme (UNDP). Unstacking global poverty: data for high-impact action. Oxford and New York; 2023.
36. Pacifico D. Estimating measures of multidimensional poverty with Stata. Stata J. 2017;17(2):687–703.
37. Parliamentary Committee for Social Affairs, United Nations Development Programme Viet Nam. Impact of HIV/AIDS on household poverty and vulnerability in Vietnam. Hanoi; 2009.
38. Pathak D, Vasishtha G, Mohanty SK. Association of multidimensional poverty and tuberculosis in India. BMC Public Health. 2021;21:2065. doi:10.1186/s12889-021-12111-4.
39. Payne CF, Houle B, Chinogurei C, Herl MCR, Kabudula CW, Kobayashi LC, et al. Differences in healthy longevity by HIV status and viral load among older South African adults. Lancet HIV. 2022 Oct;9(10):e709–e716.
40. Peña Longobardo LM, Oliva-Moreno J. Differences in labour participation between people living with HIV and the general population. PLoS One. 2018 Apr;13(4):e0195735. doi:10.1371/journal.pone.0195735.
41. Ranganathan M, Heise L, MacPhail C, Stöckl H, Silverwood RJ, Kahn K, et al. Exploring the role of transactional sex in young women’s consumption patterns in rural South Africa. Reprod Health. 2018;15:102. doi:10.1186/s12978-018-0539-y.
42. Schilling KA, Awuor AO, Rajasingham A, Moke F, Omore R, Amollo M, et al. Water, sanitation, and hygiene characteristics among HIV-positive households in rural western Kenya. Am J Trop Med Hyg. 2018 Aug;99(4):905–915. doi:10.4269/ajtmh.17-0774.
43. Sherr L, Haag K, Tomlinson M, Rudgard WE, Skeen S, Meinck F, et al. Understanding accelerators to improve SDG-related outcomes for adolescents. PLoS One. 2023 Jan;18(1):e0278020. doi:10.1371/journal.pone.0278020.
44. Tessema RA, Alemu BM. Adequacy of improved sources of drinking water, sanitation, and hygiene practice among people living with HIV/AIDS in Ethiopia. HIV AIDS (Auckl). 2021 Jan 6;13:1–11. doi:10.2147/HIV.S286976.
45. United Kingdom Health Security Agency. Positive Voices 2022 survey report. London: GOV.UK; 2024.
46. United Nations. UN80 Initiative: shifting paradigms, united to deliver. New York: United Nations; 2025.
47. United Nations Development Programme (UNDP). SDG Accelerator and Bottleneck Assessment Tool. New York: UNDP; 2017.
48. United Nations Population Fund (UNFPA). World Population Dashboard 2025. New York: UNFPA; 2025.
49. Viswanath K, Hiremath RN, Manjunath SR, Kadam DB, Raj R, Nimbannavar SM, et al. Water, sanitation, and hygiene (WaSH) practices among people living with HIV/AIDS in the era of COVID-19. J Fam Med Prim Care. 2022 Dec;11(12):5399–5403. doi:10.4103/jfmpc.jfmpc_1441_22.
50. World Bank. Poverty, Prosperity, and Planet Report 2024: Pathways Out of the Polycrisis. Washington (DC): World Bank; 2024. doi:10.1596/978-1-4648-2123-3.
51. Yates T, Lantagne D, Mintz E, Quick R. The impact of water, sanitation, and hygiene interventions on the health and well-being of people living with HIV. JAIDS. 2015 Apr;68(Suppl 3):S318–S330. doi:10.1097/QAI.0000000000000487.
52. Zhang L, Li X, Zhuang Y, Li N. World Bank aid and local multidimensional poverty in sub-Saharan Africa. Econ Model. 2022 Sep;117:106065. doi:10.1016/j.econmod.2022.106065.
